# The quantitative landscape of the neutralizing antibody response to SARS-CoV-2

**DOI:** 10.1101/2020.09.25.20201996

**Authors:** Pranesh Padmanabhan, Rajat Desikan, Narendra M. Dixit

## Abstract

Neutralizing antibodies (NAbs) appear promising interventions against SARS-CoV-2 infection. Over 100 NAbs have been identified so far and several are in clinical trials. Yet, which NAbs would be the most potent remains unclear. Here, we analysed reported *in vitro* dose-response curves (DRCs) of >70 NAbs and estimated corresponding 50% inhibitory concentrations, slope parameters, and instantaneous inhibitory potentials (*IIPs*), presenting a comprehensive quantitative landscape of NAb responses to SARS-CoV-2. NAbs with high *IIPs* are likely to be potent. To assess the applicability of the landscape *in vivo*, we analysed available DRCs of NAbs from individual patients and found that the responses closely resembled the landscape. Further, we created virtual patient plasma samples by randomly sampling NAbs from the landscape and found that they recapitulated plasma dilution assays from convalescent patients. The landscape thus offers a facile tool for benchmarking NAbs and would aid the development of NAb-based therapies for SARS-CoV-2 infection.

## Introduction

The pace of the development of neutralizing antibodies (NAbs) against the severe acute respiratory syndrome coronavirus 2 (SARS-CoV-2) has been phenomenal^1^. Over 100 monoclonal NAbs have been identified so far and several of them, namely, LY-CoV555, JS016, REGN10933/10987, VIR-7831/7832, TY027, SCTA01, BRII-196/198, CT-P59, AZD8895/1061 and MW33 are already in clinical trials^2,3^. Drugs and vaccines specifically targeting SARS-CoV-2 are not yet available. Plasma therapy, where plasma isolated from convalescent patients is injected into infected individuals, has shown some success and is in use in some countries to treat severe SARS-CoV-2 infection^4-6^. The reported NAbs have been isolated from convalescent patients and subsequently selected or engineered for improved potency^1,2^, and are therefore expected to work better than plasma therapy^7^. NAbs thus hold promise of evolving into a powerful weapon against SARS-CoV-2 infection. Passive immunization with antibodies has shown promise in several other settings, including HIV-1 infection^8,9^, autoimmune disorders^10,11^, and cancers^12,13^. With the large and rapidly growing SARS-CoV-2 NAb repertoire, a question that arises is which of these NAbs should be taken up for clinical development. A comparative evaluation of the NAbs has not been performed.

Studies identifying NAbs typically report the 50% inhibitory concentration, *IC*_50_, of the NAbs, the concentration at which viral infectivity is reduced by 50% of that in the absence of the NAbs (Fig. 1A). The inference drawn is that the lower is the *IC*_50_, the more potent is the NAb (Fig. 1A). A limitation of this approach arises from the non-linear dependence of the neutralization efficacy of NAbs on their concentrations because of which a NAb with a lower *IC*_50_ may be much less efficacious than a NAb with a higher *IC*_50_ when the two are used at physiologically relevant concentrations, which are typically much larger than *IC*_50_ (Fig. 1B). This problem was first recognized with antiretroviral drugs^14-17^. It was overcome by the construction and use of a metric called the instantaneous inhibitory potential, denoted *IIP*, which is a composite of the *IC*_50_ and the slope of the dose-response curve (DRC), *m*, the latter a measure of the extent of the non-linearity in the dependence of the efficacy on the concentration. 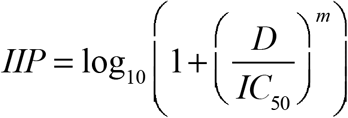 is the log decline of viremia in a single round infection assay due to the drug at concentration *D*. Thus, when two drugs are used at the same concentration, the one with the higher *IIP* would be more efficacious (Fig. 1C). Drug combinations with higher *IIP* values have been shown to have better efficacies, with HIV-1^14,15^ and hepatitis C virus (HCV)^18,19^. *IIP* has since been extended to antibodies and shown to predict the relative efficacies of HIV-1 and HCV antibodies^18,20^.

**Figure 1.**
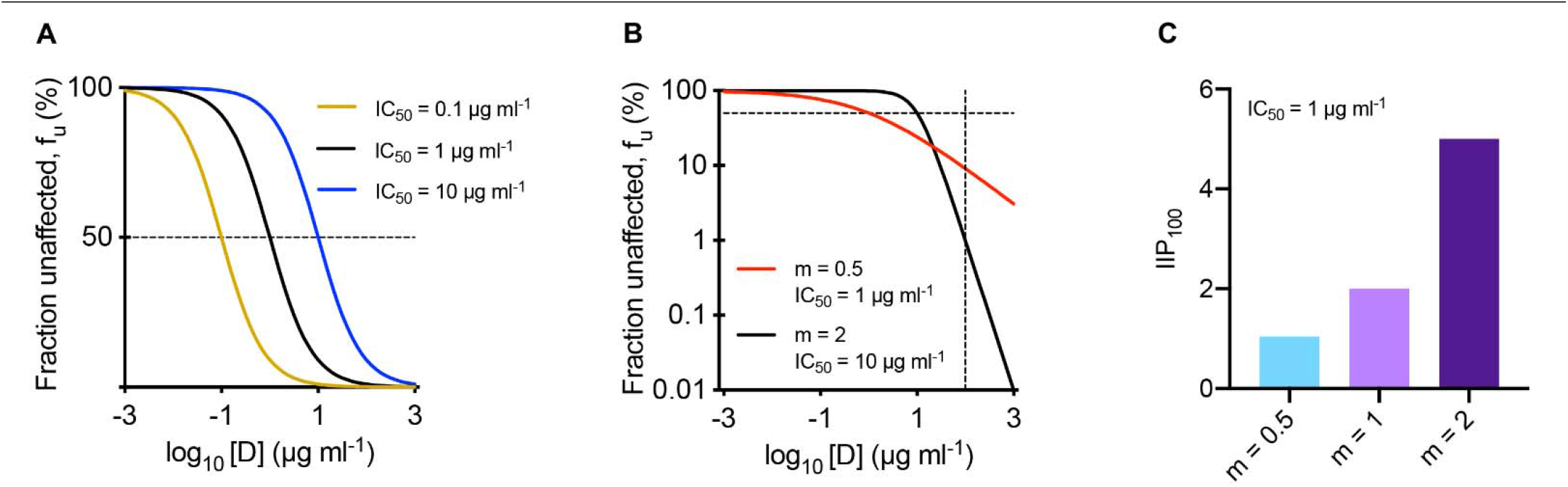
Illustration of assays and metrics characterizing NAbs. (A, B) The fraction of infection events unaffected by NAbs, *f*_*u*_, with the same *m* and different values of *IC*_50_ (A) and with different values of *m* and *IC*_50_ (B). In A and B, horizontal lines mark *f*_*u*_ = 50%. In B, vertical line marks NAb concentration, *D*, of 100 µg/ml. (C) *IIP* computed at 100 µg/ml for NAbs with the same *IC*_50_ but different values of *m*.

Here, we decided to examine whether the *IIP* could be applied to comparatively evaluate SARS-CoV-2 NAbs. Unlike *IC*_50_ values, which are routinely reported, the values of *m* have rarely been reported for SARS-CoV-2 NAbs, precluding the estimation of *IIP* for most NAbs. We therefore collated all the available *in vitro* DRCs of SARS-CoV-2 NAbs and analysed them to estimate both *IC*_50_ and *m*, and then *IIP* (Fig. 2). A comprehensive landscape of NAb responses to SARS-CoV-2 emerged. We tested the applicability of the landscape *in vivo* by examining the spectrum of responses in individual patients and by constructing virtual patient plasma samples to recapitulate plasma dilution assays.

**Figure 2.**
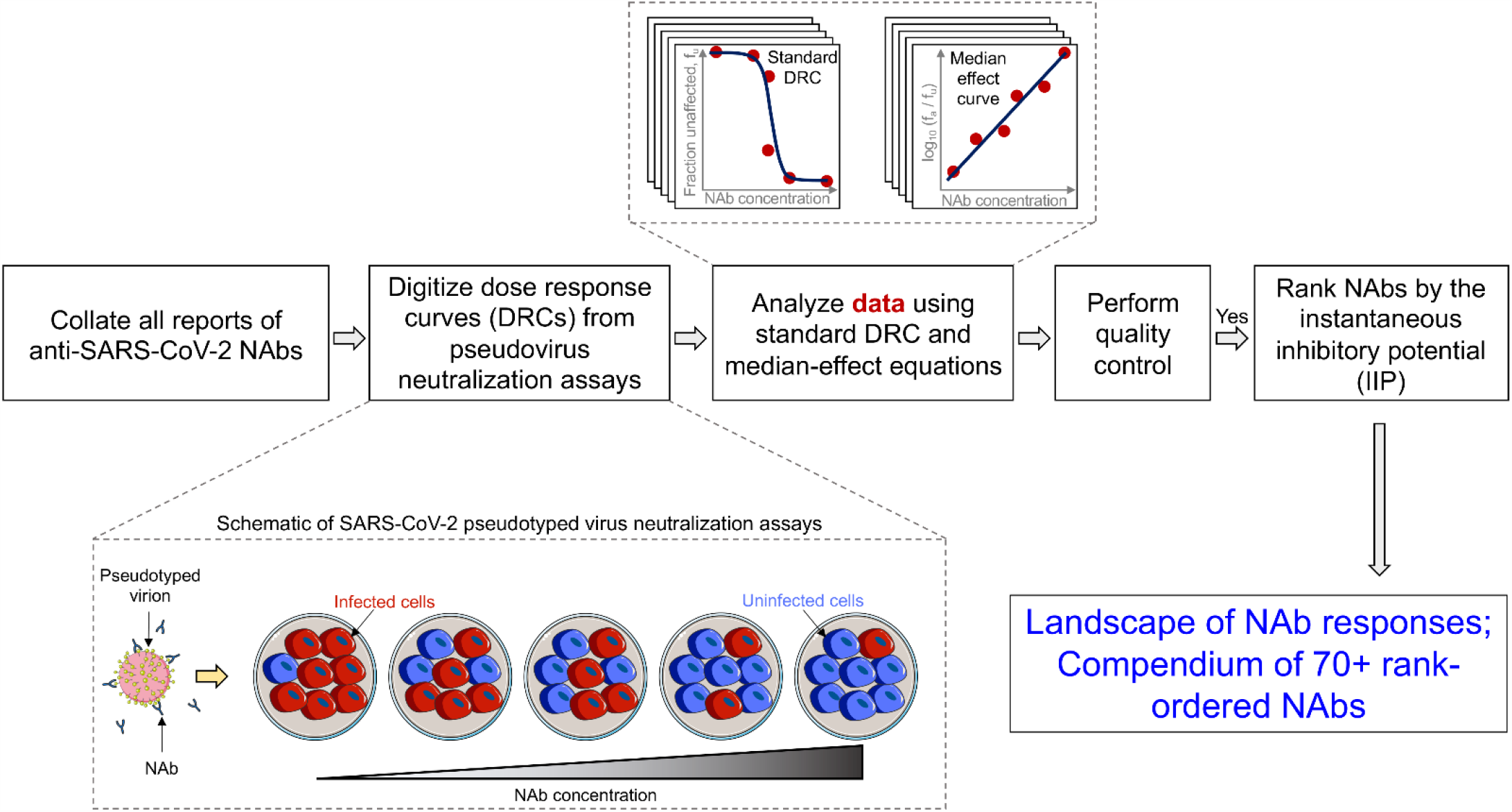
Schematic of the workflow to chart out the quantitative landscape of SARS-CoV-2 NAbs. We collated data from all studies that reported DRCs of NAbs using SARS-CoV-2 pseudotyped virions. The assays estimate the fraction of infection events affected/unaffected by the NAbs as a function of the NAb concentration. We extracted and analysed the data using both the standard DRC equation (Eq. [1]) and the median-effect equation (Eq. [2]) to estimate *IC*_50_ and *m*. We then computed 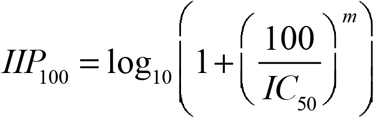 using the estimates obtained by both equations. NAbs with consistent estimates were considered for rank-ordering.

## Results

### Analysis of DRCs and the variability in IC_50_ and m

We collated and analysed the reported DRCs of over 70 NAbs obtained using SARS-CoV-2 pseudotyped virus infection assays (Figs. 2 and 3; Methods; Table S1)^21-39^. These NAbs have been proposed as the most promising from among many examined in the respective studies. A vast majority of the DRCs could be fit well with the median effect equation (Fig. 3A; Fig. S1) as well as the standard dose-response equation (Fig. 3B; Fig. S2), with the two yielding very similar estimates of the fit parameters *IC*_50_ and *m* (Table S1). The resulting estimates of *IC*_50_ were in close agreement with the reported estimates, giving us confidence in the fits (Fig. S3A; Table S1). The *IC*_50_ displayed a wide variation across NAbs, ranging from ∼10^−3^ µg/ml to ∼140 µg/ml (Fig. 3C). *m* too displayed wide variability, spanning the range ∼0.2 to 2.3 (Fig. 3D). As mentioned above, values of *m* have not been reported in previous studies. We examined whether the variability in *IC*_50_ and *m* was restricted to a particular pseudotyped virus construct or backbone used (Fig. 3F, 3G), the cell line used (Fig. 3H, 3I), or assay conditions, which could vary across studies (Fig. S3B, S3C), and found that not to be true.

**Figure 3.**
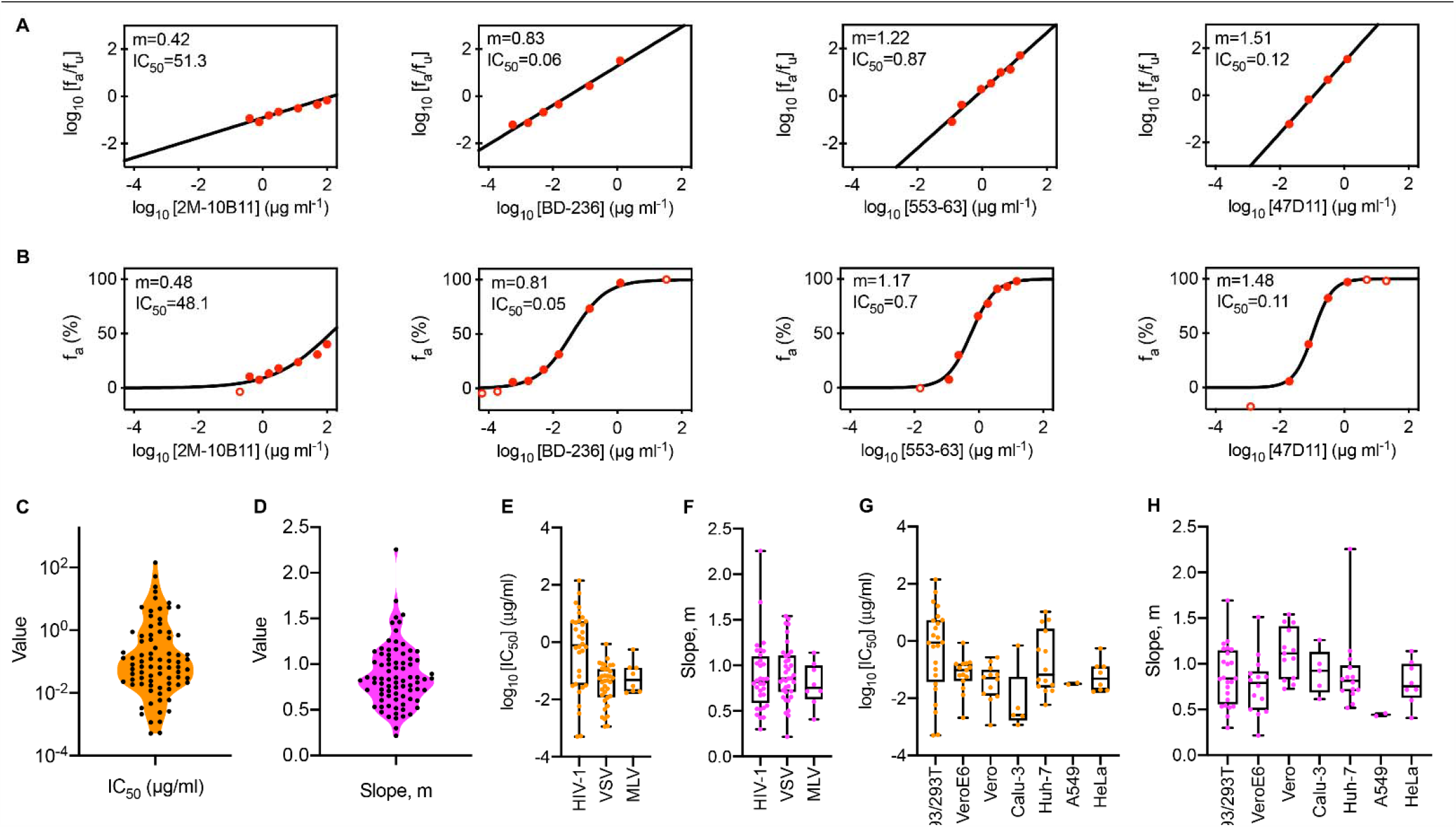
Estimates of *IC*_50_ and *m* of SARS-CoV-2 NAbs. (A, B) Fits (lines) of the median-effect equation (A) and the standard dose-response curve equation (B) to published experimental data (circles). The unit of *IC*_50_ is µg/ml. Experimental data points with 1% < *f*_*u*_ < 99% (filled circles) were considered for parameter estimation. (C, D) The best-fit estimates of *IC*_50_ (C) and *m* (D). (E-H) The variability in *IC*_50_ and *m* within different pseudotyped virus constructs or backbones (E, F) and cell lines (G, H). In G and H, 293, 293T, HeLa and A549 cells expressing ACE2 were used in the reported experiments.

The variation was thus intrinsic to the NAbs. Furthermore, akin to HIV-1 antibodies^20^, the variations in *IC*_*50*_ and *m* of the SARS-CoV-2 NAbs appeared independent. For instance, the NAbs BD-361 and REGN10954 had similar *IC*_50_ (both ∼0.04 µg/ml), but vastly different *m* (∼0.7 and ∼1.5, respectively), whereas the NAbs CC12.3 and 515-5 had vastly different *IC*_50_ (∼0.02 µg/ml and 1.6 µg/ml, respectively), but similar *m* (both ∼1). This independent variability of *IC*_*50*_ and *m* implied that *IC*_*50*_ alone was an inadequate metric to characterize the NAbs. Indeed, at concentrations of 10×*IC*_50_, REGN10954 would have an efficacy of ∼0.97 and therefore perform better than BD-361, which would have an efficacy ∼0.84, despite the two having similar values of the *IC*_*50*_.

### IIP estimates, the NAb landscape, and benchmarks

Following previous studies on HIV-1 and HCV^18-20^, we therefore computed next the *IIP* values of the NAbs at *D*=100 µg/ml. We found that the *IIP* displayed a wide range, from ∼0.3 to 7.2 (Figs. 4 and 5), giving a glimpse of the range of NAb efficacies realizable *in vivo*. (*IIP* values at *D* = 50 µg/ml displayed negligible deviations in the rank-ordering of the NAbs; Fig. S4). NAbs with high *IIPs* were those with low *IC*_50_ and high *m* (Fig. 4A). Contour lines of constant *IIP* on an *IC*_*50*_–*m* plot helped visualize the dependence of the *IIP* on *IC*_*50*_ and *m* (Fig. 4A). We found that 5 NAbs had *IIP* > 5. These, from our calculations, would be the most promising NAbs. (Similar predictions have been made with broadly neutralizing antibodies of HIV-1^20^.) The numbers increased for smaller *IIP*, with 9 NAbs between 4 and 5, 10 between 3 and 4, and so on (Fig. 4B). The highest number, 22, had modest *IIPs*, between 1 and 2. A much smaller number, 9, had *IIPs* between 0 and 1, which was predictive of poor *in vivo* efficacy.

**Figure 4.**
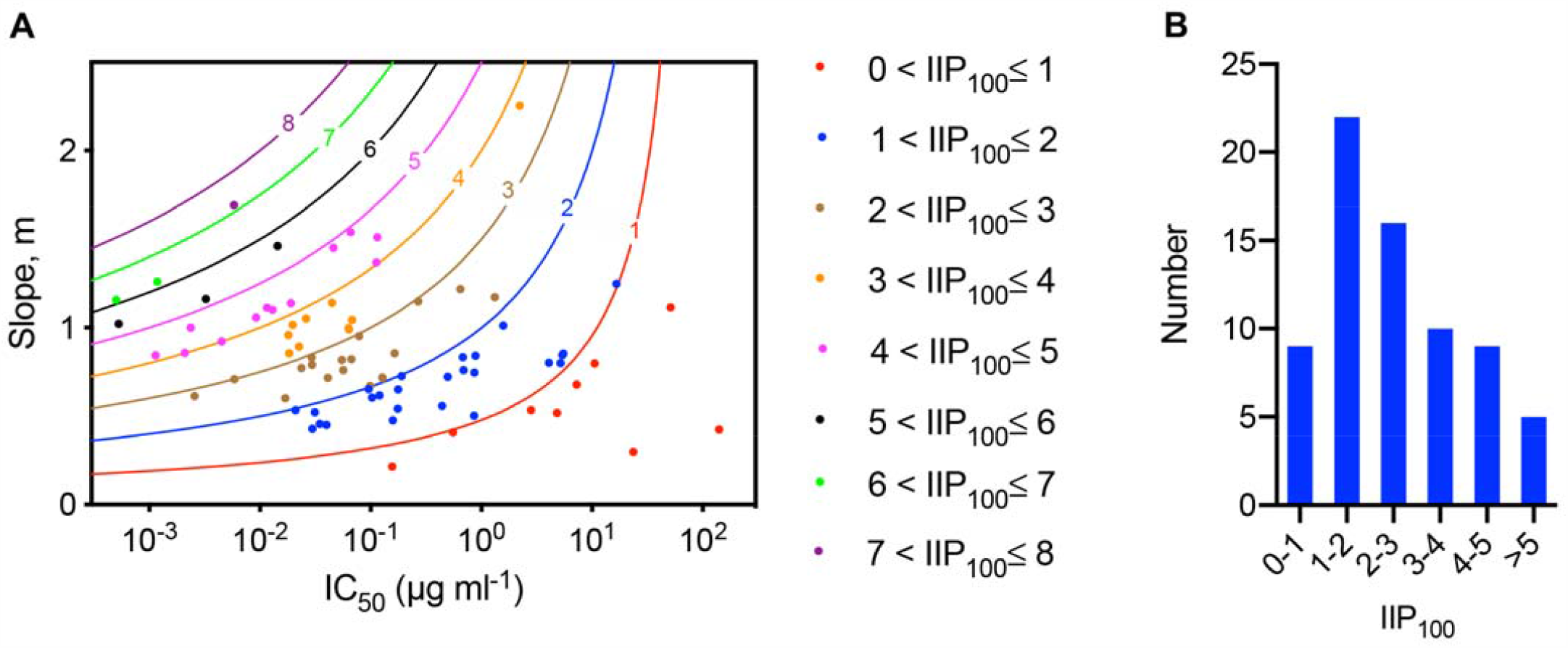
The SARS-CoV-2 NAb landscape. (A) *IC*_50_ and *m* for SARS-CoV-2 NAbs (circles colour-coded with the respective *IIP* values computed at 100 µg/ml). Each symbol represents one NAb. 8 NAbs that have multiple neutralisation curves reported are represented multiple times (Table S1). Lines are loci of points corresponding to fixed *IIP* values computed at 100 µg/ml. (B) The distribution of *IIP*_100_ values of NAbs. Average *IIP*_100_ values are used for the 8 NAbs mentioned above.

**Figure 5.**
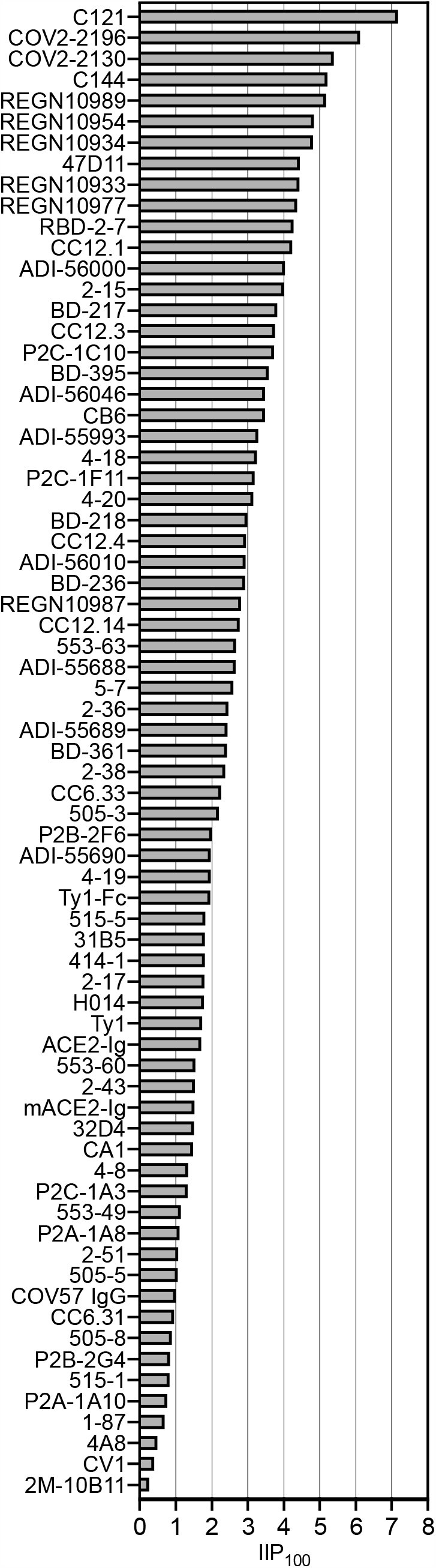
Comparison of SARS-CoV-2 NAbs based on *IIP*. A rank-order of SARS-CoV-2 NAbs based on *IIP* computed at 100 µg/ml. Average *IIP* is reported for 8 NAbs that have multiple neutralisation curves (Table S1).

This distribution of *IIP* values demonstrates the wide spectrum of neutralization efficiencies of NAbs that have all been deemed promising in different studies. The threshold *IIP* for clinical success is not known. While many NAbs with low *IIPs* may thus prove successful, it may be advantageous to choose those with high *IIPs* for they are likely to succeed with smaller dosages and/or fewer doses than those with low *IIPs*. Based on the resources available, thus, the top few NAbs, *i*.*e*., those with the highest *IIP* values (Fig. 5), could be considered for further development. We note that the top NAbs would have been different had the *IC*_50_ or *m* alone been used to characterize the NAbs, reiterating the inadequacy of these metrics individually in characterizing NAbs. (Figs. S5 and S6). The range of *IIPs* we estimated also sets benchmarks for NAbs that may be discovered/engineered in the future. They are unlikely to be competitive if they have *IIP* < 5 and more certainly so with *IIP* < 4.

### NAb responses in patients

An important question in choosing NAbs based on our analysis above is the applicability of the landscape to *in vivo* settings. The applicability of *in vitro* estimates to *in vivo* settings may not be quantitative, although proportionality has been suggested^40^. To test this here, we examined the spectrum of NAb responses reported within individual patients and compared them with the landscape. We considered reported DRCs of different NAbs from eight patients^24^ and estimated the corresponding *IC*_*50*_ and *m* (Figs. 6A, 6B, S7). We found the ranges of *IC*_*50*_ and *m* within each patient to be similar to the ranges in our landscape. For instance, in the patient COV021^24^, the *IC*_50_ range was ∼5×10^−3^ to >1 µg/ml and the *m* values were in the range ∼0.65 to 1.8. (Inhibition assays were not performed at NAb concentrations above 1 µg/ml, precluding analysis of NAbs with *IC*_50_>1 µg/ml, of which there were several^24^. Such NAbs, expected to have low *IIP* values, are therefore missing in our landscape.) Similarly, in patient COV047, the *IC*_50_ range was ∼3×10^−3^ to >1 µg/ml and the *m* values were in the range ∼0.6 to 1.5. In comparison, recall that the corresponding ranges were ∼10^−3^ to 140 µg/ml and ∼0.2 to 2.3 in the NAb landscape (Figs. 3, 4). We next computed *IIPs* of all of these NAbs and found them to lie in the range of 1 to 8, again in consonance with the landscape (Figs. 6C, 4A). These comparisons suggest that our landscape was representative of the spectrum of NAb responses within individuals.

**Figure 6.**
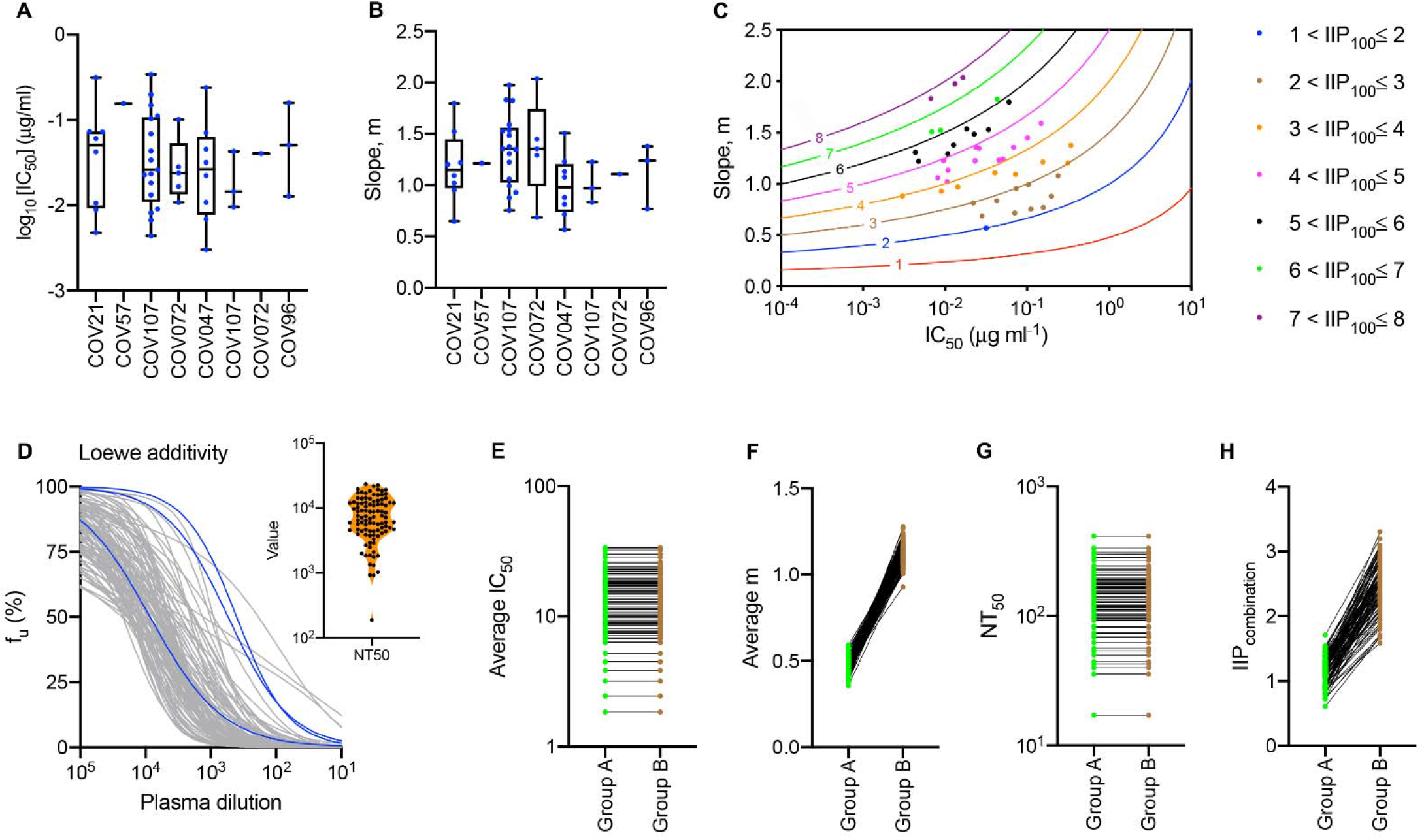
Applicability of the NAb landscape *in vivo*. (A, B) The variability in *IC*_50_ and *m* of different NAbs from eight patients (see Fig S8). The *m* values of NAbs with *IC*_50_ ≥ 1 µg/ml could not be estimated and these NAbs are not shown. (C) *IC*_50_ and *m* for SARS-CoV-2 NAbs isolated from patients (circles colour-coded with the respective IIP values computed at 100 µg/ml). Each symbol represents one NAb. Contour lines represent loci of constant *IIP*. (D) Predictions (grey lines) of the fraction of infection events unaffected by NAbs, *f*_*u*_, in the presence of increasing concentrations of plasma derived from virtual patients. We assumed ten NAbs per patient. *IC*_50_ for each NAb was sampled from the range 0.001 and 100 µg/ml and *m* from the range 0.2 and 2. Blues lines are fits to published data from three representative patients (see Fig. S8). *Inset*: half-maximal inhibitory plasma neutralizing titre, *NT*_50_, values corresponding to the dilution curves in D. (E-H) Predictions of dilution curves of hypothetical patient plasma samples. (E, F) Average *IC*_50_ (E) and *m* (F) of groups A and B. (G, H) Comparison of *NT*_50_ (G) and *IIP* (H) between groups A and B. In D-H, *D*_0_ = 100 µg/ml.

To test this further, we performed *in silico* simulations that mimic plasma dilution assays used to quantify the antiviral efficacy of convalescent patient plasma samples (Fig. 6D). We assumed that the plasma samples contained 10 different NAbs with each NAb defined by its *IC*_*50*_ and *m*. We created *in silico* virtual patient plasma samples by randomly selecting *IC*_*50*_ and *m* for each of the 10 NAbs from the ranges identified from our landscape (Fig. 4). We assumed Loewe additivity^41-43^ between the different NAbs to describe their overall efficacy. We found that with these *in silico* samples, we were able to closely recapitulate experimental serial dilution assays (Figs. 6D, S8), giving us confidence in the NAb landscape. The values of *NT*_50_, the dilution at which neutralization efficiency decreases by 50% of the undiluted plasma, we estimated (Fig. 5D inset) were also comparable to the values estimated from patient samples (∼10^1^ to 10^4^; see Ref. 24).

These comparisons also re-emphasize the need to choose NAbs or convalescent patient plasma for treatment based not only on the *IC*_50_ but also *m*. To elucidate this further, we repeated our *in silico* analysis by comparing simulated samples containing NAbs with similar *IC*_*50*_ values but low (group A) or high (group B) *m* (Fig. 6E, 6F). Although simulated plasma samples from groups A and B had similar *NT*_50_ (Fig. 6G), samples with high *m* on average had much higher values of *IIP*_100_ than those with low *m* (Fig. 6H). Thus, in interpreting plasma dilution assays and in designing plasma and NAb therapies, accounting for *m*, which has been ignored thus far, would be as important as *IC*_50_.

## Discussion

Our study presents the first quantitative landscape of the NAb responses to SARS-COV-2. We deduced the landscape by the analysis of reported data from over 70 NAbs, which we collated into an extensive and mineable compendium. Importantly, the landscape recapitulated the spectrum of NAb responses seen in convalescent patients. We also rank-ordered the NAbs based on their *IIPs*, identifying promising candidates for further development, and setting benchmarks for NAbs that may be identified in the future.

An intriguing question that emerges is the nature of the NAbs that display high *IIP* values. Correlations have been proposed between the binding affinity of antibodies for their targets and the resulting neutralization efficiency, typically *IC*_50_^44,45^. The origins of *m* are much less explored. Cooperative effects have been argued to lead to high *m*^46^. With SARS-CoV-2, these effects are yet to be elucidated. In many cases, the targets and/or the mechanism of action of the NAbs are not known. For instance, the potent NAb 47D11 targeting the spike protein S of SARS-CoV-2 blocks virus entry without preventing the binding between S and the host receptor ACE-2 required for entry^32^. As future studies establish molecular details of the SARS-CoV-2 entry process^37,47^, identifying unifying characteristics of the NAbs with high *IIP* values would become feasible, allowing rational design of even more potent NAbs.

Our analysis used data from pseudovirus assays because of the ease of interpretation of the assays and the known correlation of the *IIP* thus estimated with *in vivo* efficacy^48^. Analysis of assays using authentic SARS-CoV-2 virus, which would establish our findings on firmer footing, are not forthcoming. *m* is difficult to estimate using data from multi-round infection assays^49^. Further, viral kinetic parameters introduce confounding effects that are not readily delineated^49,50^, posing challenges that extend beyond SARS-CoV-2. Nonetheless, the ability of our NAb landscape to recapitulate patient NAb responses and plasma dilution assays suggests that our findings are likely to be consistent with the scenario *in vivo*. Our aim is not to offer accurate estimates of the *in vivo* potency of NAbs. Rather, it is to present a systematic way of comparatively evaluating NAbs for further development and a benchmark for new NAbs.

In summary, our study collates, compares, and ranks available NAbs, laying out the landscape of NAb responses currently observed including in patients, and informs ongoing efforts to develop NAb-based interventions for SARS-CoV-2 infection.

## Methods

### Data

We considered data from studies that reported dose-response curves of NAbs using SARS-CoV-2 pseudotyped virions^21-39^. The assays estimate the fraction of infection events unaffected by the NAbs as a function of the NAb concentration (Fig. 2). Data from such assays have been successfully used to evaluate *m* and *IC*_50_ of antibodies against HIV-1^20^ and HCV^18^. We extracted the data using Engauge Digitizer 12.1 and ensured consistency wherever possible with reported details, such as dilution levels used.

### Analysis of DRCs

We used both the standard dose-response curve equation (Eq. [1]) and the median-effect equation (Eq. [2]) to analyse the data.

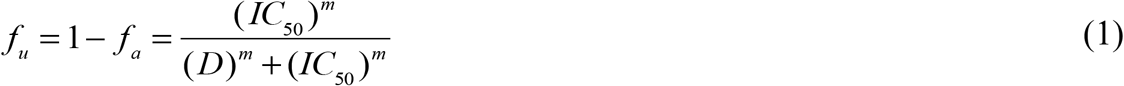

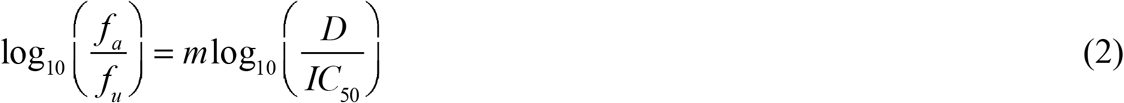

Here, *f*_*u*_ and *f*_*a*_ are the fraction of infection events unaffected and affected by the NAbs in a single round of infection, *D* is the NAb concentration, *IC*_50_ is the half-maximal inhibitory concentration and *m* is the slope. Data was fitted using the tool NLINFIT in MATLAB R2017b. Data points with 1% < *f*_*u*_ < 99% were considered for parameter estimation. We fit the data using Eq. [1] and Eq. [2] separately and obtained estimates of *IC*_50_ and *m* for each NAb as well as associated 95% confidence intervals. We then computed 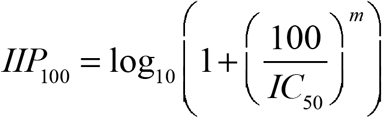 using the estimates obtained using Eq. [1] and Eq. [2]. In most cases, the *IIP*_100_ values were close to each other. We did not include NAbs for which *IIP*_100_ values estimated using the two methods deviated by 20% or more in our analysis, for the deviation indicated that such NAbs either did not conform to the trends expected by Eqs. [1] and [2] or had large uncertainties in the data precluding robust parameter estimation. We also repeated our analysis with a more liberal threshold of 30% deviation for acceptance (Fig. S9). The details of the NAbs and parameter estimates are presented in Table S1.

### In silico simulation of plasma dilution assays

We simulated plasma dilution experiments as follows. We assumed that the plasma contained *N* NAbs with equimolar concentrations. For each NAb, *IC*_50_ was sampled from the range 0.001 µg/ml to 100 µg/ml (Fig. 3C) and *m* was sampled from the range 0.2 to 2 (Fig. 3D). This range was consistent with the range seen from the spectrum of NAbs within individual patients (Fig. 6A). The reciprocal plasma dilution curve was predicted assuming Loewe additivity between the different NAbs^41,42^ using

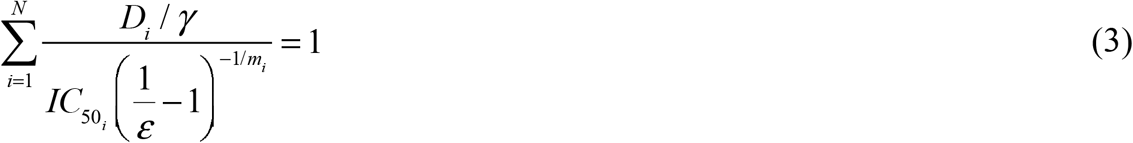

Here, γ is the plasma dilution factor. ε is the fraction of infection events affected by the plasma in a single round of infection. *D*_*i*_ is the concentration of the *i*^*th*^ NAb in the plasma before dilution, 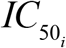 is its half-maximal inhibitory concentration and *m*_*i*_ its slope, with *i ∈* {1,2,.., *N*}. We assumed *N* = 10 in our simulations, based on the number of NAbs with significant neutralization efficacy seen in patients^25^. We estimated the value of at which 0.5 as the corresponding *NT*_50_. We chose *D*_*i*_ as *D*_0_ /N, and set *D*_0_ = 100 µg/ml.

We repeated these simulations 100 times, with each simulation representative of an individual patient. We compared the resulting predictions with observations from 3 patients, which also we digitized (Fig. S8). The equation 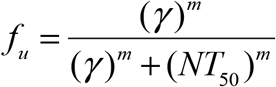 was fit to the observations from 3 patients. Here, *m* is the slope parameter, γ is the plasma dilution and *NT*_50_ is the half-maximal inhibitory plasma neutralizing titre. We also performed simulations where the *IC*_50_ values were kept similar between pairs of realizations, but *m* values were chosen from non-overlapping ranges. Using these simulations, we predicted how the expected plasma dilution assay data and the corresponding *IIP* values would vary with *m*.

## Data Availability

All data is available within the manuscript.

## Acknowledgements

This work was supported by the DBT/Wellcome Trust India Alliance Senior Fellowship IA/S/14/1/501307 to NMD.

